# Algorithmic Fairness in Computational Medicine

**DOI:** 10.1101/2022.01.16.21267299

**Authors:** Jie Xu, Yunyu Xiao, Wendy Hui Wang, Yue Ning, Elizabeth A Shenkman, Jiang Bian, Fei Wang

## Abstract

Machine learning models are increasingly adopted for facilitating clinical decision-making. However, recent research has shown that machine learning techniques may result in potential biases when making decisions for people in different subgroups, which can lead to detrimental effects on the health and well-being of vulnerable groups such as ethnic minorities. This problem, termed algorithmic bias, has been extensive studied in theoretical machine learning recently. However, how it will impact medicine and how to effectively mitigate it still remains unclear. This paper presents a comprehensive review of algorithmic fairness in the context of computational medicine, which aims at improving medicine with computational approaches. Specifically, we overview the different types of algorithmic bias, fairness quantification metrics, and bias mitigation methods, and summarize popular software libraries and tools for bias evaluation and mitigation, with the goal of providing reference and insights to researchers and practitioners in computational medicine.

## 1 Introduction

The recent years have witnessed a surge of interests on development and deployment of machine learning algorithms in healthcare. These algorithms were learned from massive health data and have demonstrated promising performance in a diverse set of problems such as skin cancer detection from lesion images^1^, prediction of the risk of acute kidney injury based on electronic health records (EHR)^2^, adaptive learning of the optimal treatment regimes for sepsis patients in intensive care^3^, and others^4^.

Despite the promise, there is growing concern that machine learning algorithms may lead to unconscious bias when making decisions against ethnic minorities, both through the algorithms themselves and the data used to learn them. For example, associations between Framingham risk factors and cardiovascular events have been shown to be significantly different across different ethnicity groups^5^. Video stream analysis algorithms for measuring the body’s spontaneous blink rate have been found to be particularly challenging for Asian individuals^6, 7^. Undiagnosed silent hypoxemia, which can be detected by pulse oximetry using light to monitor vital signs, occurred approximately three times more frequently in Black people due to the fact that dark skin responds differently to those light wavelengths ^8^. In these cases, the software system may bring in additional or exacerbate health equity issues^7^.

With machine learning models gaining more and more attentions in medicine, it is crucial to be aware of the potential related bias and disparities, understand their causes, and mitigate them. This review will help achieve this goal by providing an overview of the existing literature studying the sources of bias and disparities in computational medicine, their quantification metrics, and mitigation strategies. We will also summarize outstanding questions and point out future directions.The PRISMA diagram of the literature reviewed in this paper is shown in Fig. 1.

**Figure 1.**
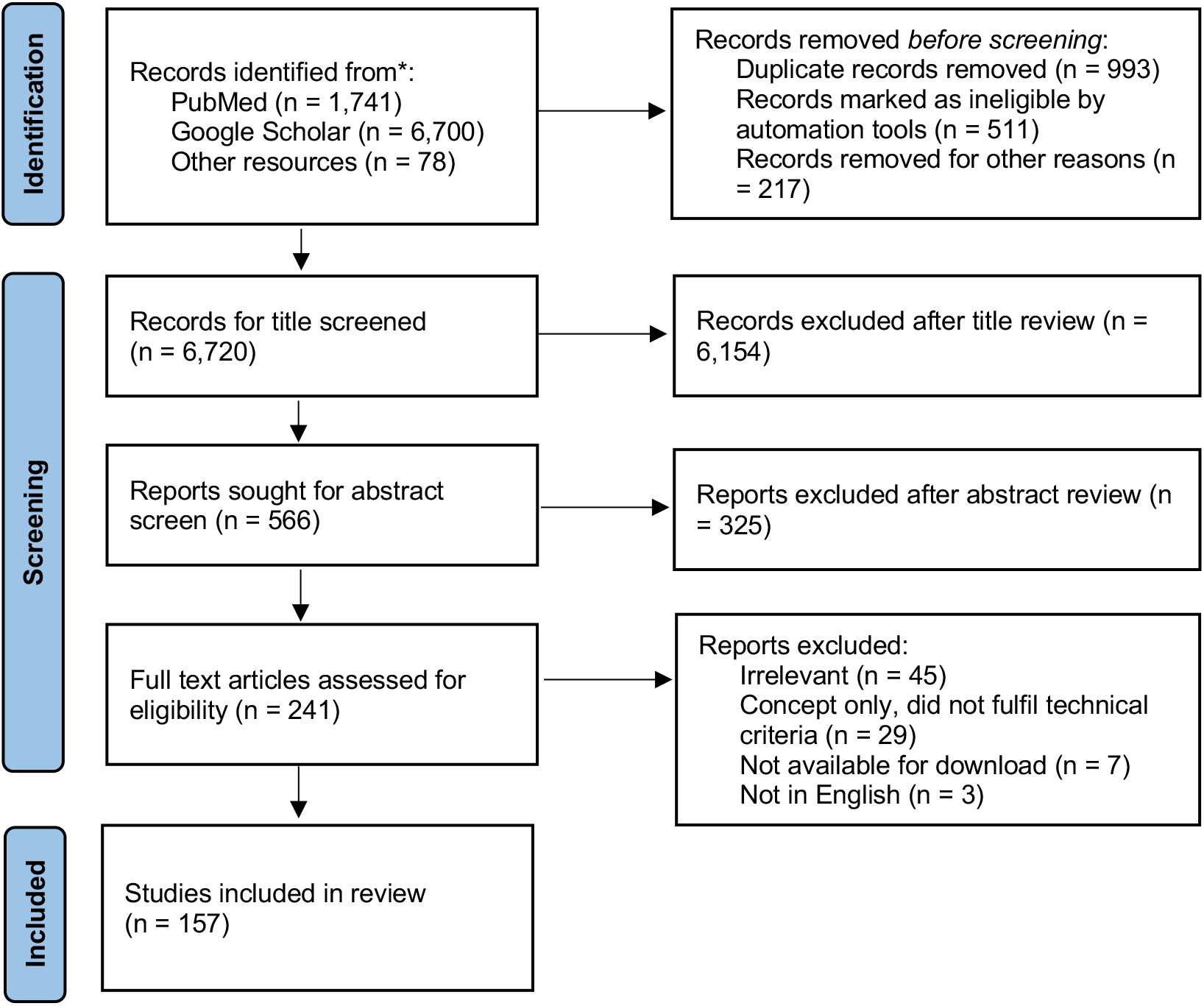
PRISMA flow diagram: disparity and fairness in computational medicine.

### Difference with Existing Reviews

Mehrabi et al.^9^ built a taxonomy of machine learning related fairness in different real world application contexts. Rajkomar et al.^10^ introduced the principles of distributive justice and provided guidance to clinicians on how to prioritize each principle when facing with potential bias in model development and deployment. Gianfrancescogian et al.^11^ summarized the potential bias sources for electronic health records (EHRs) and provided recommendations on appropriately mitigating them. Fletcher et al.^12^ described three basic criteria (*i.e*., Appropriateness, Fairness, and Bias) for evaluating machine learning and AI systems in the context of global health. Mhasawade et al.^13^ focused on the interactions among different cultural, social, and environmental factors and their impact on individual and community health, how they will impact the fairness of machine learning algorithms and how machine learning, public and population health can work together to achieve health equity. Different from these existing works, this review summarizes sources and quantification methods for bias in computational medicine and how they will impact downstream machine learning models, as well as potential strategies to mitigate them through computational algorithms.

## 2 Computational Bias

We categorize computational biases into three different types according to their sources, *data bias, measurement bias*, and *algorithm bias*. We will introduce them in this section and provide examples in medical context.

### 2.1 Data Bias

Machine learning algorithms are all learned from data sets^14^. For example, classification models try to accurately map the sample input features to a set of pre-specified classes based on the observations from a set of training data. Clustering models aim at identifying grouping structures of a given data set. In this case, if the data set is over or under representing certain sample groups, the machine learning models learned from the data will be biased. For instance, studies found that patients of low socioeconomic status may have limited access to health care^15, 16^. Consequently, compared to patients with better socioeconomic status, these patients may have less information in their EHRs or imaging and thus underrepresented in the data from which a machine learning model will be learned. This will lead to poorer model performance on this particular patient group. Below we list potential sources of data bias in medicine.

#### 2.1.1 Sample bias

Sample bias, also known as selection bias, occurs when the selected data can not represent the real environment in which a model will be deployed^17^. For example, melanoma detection algorithms based on classification of skin lesion images^1^ may perform poorly on colored skins if the training images are mostly with white skin^18^. For the same reason, Face2Gene, a machine learning algorithm to recognize Down syndrome based on facial images, performed much better in Caucasian (accuracy 80%) than in African (accuracy 36.8%)^19^.

#### 2.1.2 Allocation Bias

Allocation bias is relevant to clinical trials of interventions, which arises if there are systematic differences in how participants are allocated to treatment and control groups^20^. If researchers know or are able to predict which participants would benefit from an intervention, it would affect how they recruit participants and how they assign them to different groups so that they can select subjects with a good prognosis for trials. Allocation concealment could protect the randomization process, keep participants unaware of the intervention to be assigned before entering the study, and prevent prediction of subsequent allocations in actual clinical trials^20^. Recently there were studies trying to emulate clinical trials with real world data such as EHRs^21, 22^. In this case, allocation bias could exist as the treatment and control groups are already observed in the data. This can lead to potentially bias estimations of treatment effects with machine learning models^23^.

#### 2.1.3 Attrition Bias

Attrition bias can occur if there are systematic differences in the way participants are dropped from the study, as different rates of losses to follow-up in the exposure groups may alter the characteristics of these groups^20^. Attrition bias will be more severe in observational data analysis as patients may move to another place or be transferred to another hospital, which will impact the machine learning model aiming at prediction of clinical events.

#### 2.1.4 Publication bias

Publication bias occurs when a study is published and not depending on its own results^24^. Empirical studies consistently show that studies with positive or statistically significant results are easier and take less time to be published than studies without significant results^25, 26^. This can make it difficult for decision makers to distinguish between sound evidence and overestimate the effectiveness of treatment or models^26^. For example, since the start of the COVID-19 pandemic, studies on COVID-19 is being published at a rapid rate. However, many peer-reviewed publications were with a limited number of patients included and showed a high risk of bias ^27^.

### 2.2 Measurement bias

Measurement bias is a systematic error that occurs when the data are labeled inconsistently, or study variables (*e.g*., disease, exposure) are collected or measured inaccurately^28^. A recent example is there is a large disparity in the quality of COVID-19 data reporting across India^29^. Below we list several common causes of measurement bias.

#### 2.2.1 Response bias

Response bias usually occurs in survey-based studies. When respondents tend to give inaccurate or even wrong answers on self-reported questions, the survey results will be affected^30^. For instance, people tend to paint the best picture of themselves, or feel pressured to provide socially acceptable answers^31^. In addition, misleading questions can lead to biased answers. Respondents may not have realized they weren’t giving the answers in the way the investigator wanted them to^30^. In addition, people who are willing to answer survey questions are often different from those who are not^32^. Consequently, this will impact the machine learning algorithms trained on surveys or patient reported outcomes.

#### 2.2.2 Recall Bias

Recall bias usually occurs during the data annotation phase of a project^33^. This happens when similar data are inconsistently labeled, thus leading to low accuracy. A participant may erroneously provide responses that rely on his/her ability to recall past events. However, recalling events of interest that happened long ago can be particularly difficult. For example, a publicity related to association between measles, mumps and rubella (MMR) vaccine and autism^34^ influenced how often parents of autistic children recalled their child’s regressive symptoms^35^. This may lead to the observation of a completely or partially untrue association between MMR and autism, which would subsequently impact the algorithms for inferring such associations.

#### 2.2.3 Observer Bias

The observer bias occurs when the methods or procedures used to observe and record information for research leading to a systematic deviation from the facts^36^, due to bad habits or lack of training in using measuring equipment or data sources^37^. Although some results of the diagnostic studies and physical examinations are objective, the symptoms and most findings of medical examinations are more or less subjective and prone to observer bias^38, 39^. Hrobjartsson *et al*.^40^ provided empirical evidence for observer bias in randomized clinical trials with results that involved subjective measurement scales. Consequently, these bias will be carried on to the machine learning models trained from these data.

### 2.3 Algorithm Bias

Another source of bias is from the algorithms themselves^41^, which can be algorithm specific or agnostic. Algorithm specific bias is linked to their intrinsic hypotheses^42^. For example, logistic regression models assume the relationships between input and target variables are linear, but this may not be the reality. This bias makes the algorithm challenging to capture the actual input-output relationships in the data. We also list two types of algorithm agnostic bias as below.

#### 2.3.1 Loss Function Bias

The loss function measures the difference between the output produced by the machine learning algorithm and the ground truth outcomes. It is used to evaluate how well the machine learning algorithm fits the data. Typical machine learning algorithms attempt to minimize such prediction loss on the training data, which is typically measured by adding up all prediction losses on individual samples. However, if certain group of samples are more representative (e.g., white patients in a population^43^), the corresponding model will be better trained for this group.

#### 2.3.2 Post-hoc Confirmation Bias

Although many machine learning models have been developed for binary classifications (e.g., disease diagnosis), they typically generated continues prediction scores and a cutoff threshold was needed for dichotomizing the predictions, and its optimal value is typically determined with post-hoc data driven analysis. The choice of cut points can introduce bias to diagnostic research. For example, Ewald analyzed the simulated data sets of test results from subjects with or without a particular disease and found that the use of data-driven cut points exaggerated test performance in many cases^44^.

## 3 Fairness Metrics

The previous section has summarized the various potential sources of computational bias. Another important question is how can we quantify such bias given a specific healthcare context or data set. In this section, we will review the various bias quantification measures, which are referred to as fairness metrics. Mathematical notations that are used in this section are summarized in Table 1.

**Table 1.** Notations and Symbols.

| Symbol | Description |
| --- | --- |
| $A \in \{0, 1\}$ | Binary protected attribute |
| $X \in \mathbb{R}^d$ | Other observable attributes |
| $U$ | Relevant latent attributes not observed |
| $Y \in \{0, 1\}$ | The outcome to be predicted |
| $\hat{Y} := f(X, A) \in \{0, 1\}$ | The prediction of $Y$ |
| $\hat{Y}_{A \leftarrow a}$ | Counterfactual value, *i.e.*, what would $\hat{Y}$ have been if $A$ had been equal to $a$ |

To facilitate the description of fairness metrics, we begin by a case study to build an alerting algorithm in ICU setting (e.g., for developing sepsis^45^) with machine learning based on EHR, and race (*i.e*., Black and White is considered) as the protected attribute, which means we want to quantitatively examine if the algorithm behaves differently for black and white patients using various fairness metrics^46, 47^.

### 3.1 Fairness through unawareness

*Fairness through unawareness* requires to not include the protected attribute (*e.g*., race in our case study) as an independent variable in the model^51–53^. This method has been shown to be ineffective in situations where there are highly relevant features (*e.g*., proxies for protected attributes). For example, race may be related to zip code, socioeconomic status or disease predisposition. Therefore, simply removing protected attribute is not enough.

### 3.2 Demographic parity

*Demographic parity*, also known as *statistical parity* or *independence*, requires that the overall proportion of individuals in a protected group predicted as positive (or negative) to be the same as that of the overall population^49^. Although it is intuitive to understand, prior studies^54^ found that optimizing demographic parity may prevent the model from taking into account relevant clinical characteristics related to protected variables and outcomes, thereby reducing the performance of the model for all groups.

### 3.3 Equalized Odds

Unlike demographic parity, *equalized odds*^50^ *allows the prediction Ŷ* to depend on protected attribute *A*, but only through the target variable *Y*. This encourages the use of features that are directly related to *Y*, rather than through *A*^50^. *To achieve equalized odds, both true positive rates (TPR) and true negative rates (TNR) of all groups defined by A* are equal up to a fixed tolerance *T*. Compared to demographic parity, equalized odds is more flexible as it does not prevent learning a predictor where there is a real association between the protected attribute and the outcome^54^.

### 3.4 Equal Opportunity

*Equal opportunity* checks whether the positive label is equally and accurately predicted by classifier for all values of the protected attribute^50^. In contrast to *equalized odds*, it is stronger because it means that all possible thresholds are equally likely to be met and therefore requires that all groups get the same ROC curve, but the decision threshold can be adjusted to satisfy *equalized odds*^54^.

### 3.5 Individual Fairness

At a high level, individual fairness requires that any two individuals who are similar in the context of a given task should be treated similarly^51, 55^. Clearly, individual fairness is more strict than group fairness defined by the protected attribute. The practical use of this concept is often limited due to the challenges of defining a appropriate similarity metric to encode the desired concept of fairness^51, 54^. In addition, there were also arguments that individual fairness is an inadequate as similar treatment is not enough to achieve fairness, thus it shouldn’t be used alone to detect bias or evaluate whether algorithms are fair^56^.

### 3.6 Counterfactual Measures

Counterfactual fairness is a potential way to explain why bias occurs. It states that a model is fair if its predictions about a particular individual in the real world is the same as it is in the counterfactual world, *i.e*. if the individuals is in a different protected group^48^. We list the mathematical definition of counterfactual fairness in the last row of Table 2, where *Ŷ*_*A←a*_ represents the the prediction *Ŷ* if *A* had taken value *a*. This metric considers the predictor to be fair if its prediction remains unchanged when the protected attribute of each sample is flipped to its counterfactual value. A close concept of counterfactual fairness is counterfactual reasoning^57^. Some studies have shown that counterfactual reasoning is susceptible to biases such as outcome bias (that is, evaluating the quality of decisions when the outcome is known)^58^. In addition, it has been suggested that counterfactual reasoning may negatively affect the process of causality identification^59^. These concerns raise questions about the practical applicability of counterfactual measures.

**Table 2.** Summary of Fairness Metrics

| Type | Definition | In our case study |
| --- | --- | --- |
| Fairness Through Unawareness<sup>48</sup> | No protected attribute $A$ is explicitly used in the decision-making process: $\hat{Y} = f(X, A) = f(X)$ | Train the model without race variable |
| Demographic Parity<sup>49</sup> / Statistical Parity / Independence | The outcomes must be equal: $\mathbb{P}(\hat{Y}\vert A = 0) = \mathbb{P}(\hat{Y}\vert A = 1)$ | Blacks and Whites developed to sepsis at equal rates |
| Equalized Odds<sup>50</sup> / Separation | Different groups deal with similar odds, if $\hat{Y}$ and $A$ are independent conditional on $Y$: $\mathbb{P}(\hat{Y} = 1\vert A = 0, Y = y) = \mathbb{P}(\hat{Y} = 1\vert A = 1, Y = y), y \in \{0, 1\}$ | The true positive rates (of those who actually developed sepsis, how many were correctly predicted to be positive) and false positive rates in Blacks and Whites are equal |
| Equal Opportunity<sup>50</sup> | The true positive rates in the unprivileged group and privileged group are equal. $\mathbb{P}\{\hat{Y} = 1\vert A = 0, Y = 1\} = \mathbb{P}\{\hat{Y} = 1\vert A = 1, Y = 1\}$ | The true positive rates in Blacks and Whites are equal |
| Individual Fairness<sup>51</sup> | Similar individuals have similar predictions. Formally, given a metric $d(\cdot, \cdot)$, if individuals $i$ and $j$ are similar under this metric (*i.e.*, $d(i, j)$ is small), then their predictions should be similar: $\hat{Y}(X^{(i)}, A^{(i)}) \approx \hat{Y}(X^{(j)}, A^{(j)})$. | Similar patients have similar chance to develop sepsis |
| Counterfactual Fairness<sup>48</sup> | Predictor $\hat{Y}$ is counterfactually fair if under any context $X = x$ and $A = a$, $Pr(\hat{Y}_{A \leftarrow a}(U) = y\vert X = x, A = a) = Pr(\hat{Y}_{A \leftarrow a'}(U) = y\vert X = x, A = a)$, for all $y$ and for any value $a'$ attainable by $A$ | The predicted outcome does not change if the values of the sensitive variables change |

Different metrics have different characteristics, and these aforementioned fairness metrics cannot be achieved at the same time, except in highly restricted special cases^60^. Both equalized odds and demographic parity focus on group fairness. Although their calculations and reasoning are simple and intuitive, the derived models may be discriminatory to structured subgroups with protected attributes, leading to fairness gerrymandering^54, 61^. The concept of individual fairness potentially alleviate the issues of group fairness metrics by forcing any two individuals who are similar at a given task should be similarly classified. However, it is challenging to a domain-specific similarity measure, thus the practical use of individual fairness is often limited. Clinical prediction models may produce unfair results based on particular metrics. There is no clear consensus on what metric should be used in each scenario, researchers should choose the fairness metrics based on the given context.

## 4 Bias Mitigation

With the various sources of bias and different fairness metrics, in this section we will summarize different bias mitigation approaches for achieving algorithmic fairness. These methods can be categorized as pre-processing^62^, in-processing^63–66^, and post-processing methods^67^, which are detailed below.

### 4.1 Pre-processing

Data pre-processing refers to the procedures of cleaning and preparing raw data for building machine learning models^68^. Pre-processing methods can potentially remove the bias from the data.

#### 4.1.1 Sampling

Sampling is a popular preprocessing method to ensure the datasets are balanced across different groups^69^. If the data set is large, the majority group can be randomly sampled to the same size as the minority group without much information loss. However, if there is no redundant data, it is more common to oversample minority groups. Popular algorithms, like synthetic minority oversampling technique (SMOTE)^70^ or its variations, such as SMOTE-ENC^71^, Borderline-SMOTE^72^. However, healthcare data (such as EHRs or questionnaires) are typically complicated, and are thus challenging to be synthesized without overfitting^12^. In addition to sampling, collecting more data with good planning is also important to mitigate potential bias more objectively ^43^.

#### 4.1.2 Reweighting

Reweighting is to impose different weights on each group-class combination based on the conditional probability of class by protected attribute, so that the protected attribute is independent of the outcome^62^. As a representative method, inverse propensity score weighting (IPW)^73^ is often adopted to adjust poorly sampled data. It involves estimating the probability of individual participants in particular groups and then analysing the re-weighted samples of these participants^74^. However, IPW adjusts the distributions of all variables simultaneously, which may potentially increases imbalances and bias^75^. Borland *et al*.,^76^ presented dynamic reweighting (DR) to correct selection bias with interactive visual analysis.

### 4.2 In-processing

In-processing methods aims at developing unbiased models directly from the data. A straightforward approach to achieve this goal is to remove the protected attribute from the model as we introduced in Section 3.1. However, if there are dependencies between the protected attribute and other covariates, the information of the sensitive attributes will “leak” into the decision.

#### 4.2.1 Prejudice Remover

Prejudice refers to the fact that there is statistical dependence between the protected attribute and the predicted outcome or other independent variables^77^. Prejudice remover aims at learning a predictor whose predictions are independent of the protected attribute. For example, Kamiran and Calders *et al*.^78^ proposed the concept of discrimination-aware classification and developed an algorithm to “clear away” such dependencies by “massaging the data” before applying traditional classification algorithms. Calders and Verwer ^63^ proposed a discrimination-free naive-Bayes through post-hoc processing, independent model training and balancing across different protected groups, or latent variable modeling. Kamishima et al.^65^ proposed a prejudice remover regularization to enforce the prediction’s independence on the protected attribute. Zafar et al.^64^ proposed the concept of “disparate mistreatment” as different misclassification rates across different protected groups, and introduced a measure for decision boundary based classifiers, which further can be incorporated into the classifier optimization objectives as constraints to remove prejudice. With more and more machine learning models being developed for clinical risk prediction, there has been intense discussions on the ethical concerns^79, 80^. These prejudice remover approaches can potentially make these algorithms fair.

#### 4.2.2 Adversarial Learning

Adversarial learning^81^ is a learning paradigm originally designed for generating fake samples to confuse the model. Typically there is a generator guaranteeing the generated fake samples which are close to real samples, and a discriminator to discriminate the fake samples from the real ones. The goal of adversarial learning is to learn a generator to generate samples that the discriminator cannot really tell they faked or no. Pfohl *et al*.^54^ applied adversarial learning for developing an “equitable” risk prediction model for atherosclerotic cardiovascular disease (ASCVD) with EHR. They used the generator to build the risk predictor and discriminator to enforce equalized odds for the predicted risks across different protected groups.

#### 4.2.3 Other Learning Strategies

Another closely related topic is interpretable learning^82^, as interpretable models can allow the decision makers to better understand why certain predictions are made and make necessary modifications. Recent work at the FICO Data Science Lab has shown that interpretable neural networks can help uncover and eliminate data biases in models. Even in cases where the data is deliberately biased toward one subset of the population over another, the method minimizes the pickup of signals that are biased toward the core relationship^83^. Similar argument has also been made by Rudin^84^ that interpretable models are more preferred in high stakes decision making scenarios such as healthcare than black-box models.

Independent learning is another bias mitigation strategy which trains a machine learning model for each protected group^85^. However, this may sacrifice the training data sample sizes and reduce the model performance^85^. Gao and Cui^85^ introduced a transfer learning approach to align the sample distributions across different protected groups. They demonstrated their method can achieve improved performance in underrepresented groups and effectively reduce disparity with cancer multiomics data.

### 4.3 Post-processing

The post-processing approach treats off-the-shelf predictors as black boxes and achieves fairness through adjustment of their predictions. For example, Hardt et al. ^50^ proposed equalized odds post-processing and calibrated equalization odds post-processing, which aims to solve for the probabilities of changing output labels to achieve the equalized odds objective. Kallus *et al*.^86^ proposed to adjust the risk scores of the instances in the disadvantaged group with a parameterized monotonically increasing function to minimize the performance disparity. Cui et al.^87^ proposed to adjust the ranking order of the samples across different protected groups according to their predicted scores with a dynamic programming procedure to achieve fairness without sacrificing prediction accuracy. One practical challenge for post-processing methods is that the involved adjustments are typically not explainable. Pan et al.^88^ proposed a causal analysis approach that can quantitatively attribute algorithm performance disparity onto different causal decision paths, so that the paths with large contributions can be removed as post-processing.

In practice, these three types of methods work at different stages of a machine learning pipeline: pre-processing manipulates the data through sampling or weighting before building the model, in-processing enforces fairness constraints during model building, and post-processing makes adjustments after the model was built. Different strategies have different assumptions, therefore it is challenging to have a golden standard. A recent research from Park et al.^89^ compared different risk mitigation methods in the context of postpartum risk prediction and found that these methods could indeed reduce bias but different methods can lead to different results. Therefore the practitioners should try to test different approaches and evaluate their impact in the particular context they were applied to.

## 5 Popular Software Libraries

We summarize existing popular algorithmic fairness research software libraries in Table 3.

**Table 3.** Popular library for fairness research

| Project Name | Developer | Description |
| --- | --- | --- |
| FairMLHealth<sup>90</sup> | KenSci | Tools and tutorials for evaluating bias in healthcare machine learning. |
| AIF360<sup>91</sup> | IBM | Fairness metrics for datasets and machine learning algorithms, interpretation of the metrics, and approaches for reducing bias in datasets and models. It is available in both Python and R. |
| Fairlearn<sup>92</sup> | Microsoft | A Python package to evaluate fairness and mitigate any observed inequities. Fairlearn includes mitigation algorithms and metrics for model evaluation. It also contains Jupyter notebooks with examples of Fairlearn usage. |
| Fairness-comparison<sup>93</sup> | Sorelle *et al.* | Compare fairness-aware machine learning techniques. It aims to facilitate benchmarking of fairness-aware machine learning algorithms. |
| MEASURES<sup>94</sup> | Cardoso *et al.* | A benchmark framework for assessing discrimination-aware models. |
| Fairness Indicators<sup>95</sup> | Google | A suite of tools built on top of TensorFlow Model Analysis that enable regular evaluation of fairness metrics in product pipelines. |
| ML-fairness-gym<sup>96</sup> | Google | A general framework for studying and exploring long-term equity effects in carefully constructed simulation scenarios where learning subjects interact with the environment over time. |
| themis-ml<sup>97</sup> | Niels Bantilan | A Python library built on top of pandas and sklearn that implements fairness-aware machine learning algorithms. |
| FairML<sup>98</sup> | Julius Adebayo | A Python toolkit for auditing machine learning model deviations. |

## 6 Conclusions

In this review, we summarized the current research on algorithmic fairness in computational medicine. We first described the three types of computational bias: data bias, measurement bias, and model bias. Then we presented the fairness quantification metrics that are used in various literature. Additionally, we introduced three types of bias mitigation methods, namely, pre-processing, in-processing and post-processing, and listed the popular software libraries and tools for bias evaluation and mitigation. Fairness is not just the result of rigorous and thoughtful research, but rather the social and political processes needed to advance health equity^99^. With machine learning and artificial intelligence models gaining more and more attentions, we should be aware of these issues when designing the models and appropriately mitigate them. To help achieve this goal, we further list some probably encountered directions or open questions in Table 4.

**Table 4.** Open questions and future directions.

| Open Questions | Description |
| --- | --- |
| Data collection | As data is the source for building machine learning models, it is critical to be aware of the potential bias and improve the diversity and inclusiveness during the data collection process. |
| Multiform fairness | Different types of fairness are sometimes incompatible. For example, a model could be fair for equal positive and negative predictive values, but unfair for equalized odds (and vice versa). It is important to understand which types of fairness are achievable under which scenarios. |
| Algorithm explainability | Explainable models can reveal how a machine learning algorithm works and thus potentially alleviate decision bias. However, on the other hand, interacting with incorrect recommendations paired with explanations that contain limited but easily interpretable information can adversely affect the clinician's treatment choices<sup>100</sup>. Understanding such interaction between algorithm explainability and bias is important for medical machine learning. |
| Model generalization | Fairness in machine learning goes beyond preventing models from harming protected populations. It can also help focus care where it is really needed. The data used to develop the model may not be generalized to the data used during the deployment of the model (training-serving skew)<sup>10</sup>. Thus, besides model design and evaluation, fairness should also be incorporated into the scenario where the model is going to be deployed<sup>101</sup>. |

## Data Availability

All data produced in the present work are contained in the manuscript

## Contributors

All authors read and approved the final version of the manuscript. JX drafted the manuscript. FW made thorough revisions to the draft. YX performed an initial literature review on computational bias. WW, YN and ES performed the literature review and data abstraction on bias mitigation methods. All authors contributed to the writing and editing of the manuscript. JB and FW conceived the idea.

## Acknowledgements

FW is supported by NSF 1750326, NIH R01AG076234, R01MH124740 and RF1AG072449. YX is supported by CORONAVIRUSHUB-S-21-00188. YN is supported by NSF 1948432 and 2047843. WHW is supported by NSF 2029038 and 2135988. JB is supported by NIH R01AG076234, R01CA246418, R21CA253394, R21AG068717, and R21CA245858.

## Declaration of Competing Interests

None declared.

## Notes

### Competing Interest Statement

The authors have declared no competing interest.

### Funding Statement

This study did not receive any funding

